# COVID-19 in Youth Soccer

**DOI:** 10.1101/2020.09.25.20201616

**Authors:** Andrew M. Watson, Kristin Haraldsdottir, Kevin Biese, Leslie Goodavish, Bethany Stevens, Timothy McGuine

## Abstract

**Purpose:** The purpose of this study was to determine the case and incidence rates of COVID-19 among youth soccer players and evaluate the relationship with background COVID-19 risk and phase of return to play.

**Methods:** Surveys were distributed to soccer clubs throughout the country regarding their phase of return to soccer (individual only, group non-contact, group contact) and date of reinitiation, number of players, cases of COVID-19, and risk reduction procedures that were being implemented. Overall case and incidence rates were compared to national pediatric data and county data from the prior 10 weeks where available. Finally, a negative binomial regression model was developed to predict club COVID-19 cases with local incidence rate and phase of return as covariates and the log of club player-days as an offset.

**Results:** 129 clubs responded, of whom 124 had reinitiated soccer, representing 91,007 players with a median duration of 73 days (IQR: 53-83 days) since restarting. Of the 119 that had progressed to group activities, 218 cases of COVID-19 were reported among 85,861 players. Youth soccer players had a lower case rate and incidence rate than the national rate for children in the US (254 v. 477 cases per 100,000; IRR = 0.511, 95% CI = [0.40-0.57], p<0.001) and the general population from the counties in which soccer clubs were based where data was available (268 v. 864 cases per 100,000; IRR = 0.202 [0.19-0.21], p<0.001). After adjusting for local COVID-19 incidence, there was no relationship between club COVID-19 incidence and phase of return (non-contact: β=0.35±0.67, p=0.61; contact: β=0.18±0.67, p=0.79). No cases were reported to have resulted in hospitalization or death. 100% of clubs reported having a plan in place to reduce the risk of COVID-19 and utilizing multiple different risk reduction procedures (median 8, IQR 6-10).

**Conclusions:** The incidence of COVID-19 among youth soccer athletes is relatively low when compared to the background incidence among children in the United States and the local general population. No relationship was identified between club COVID-19 incidence and phase of return to soccer. Youth soccer clubs universally report implementing a number of risk reduction procedures.

## INTRODUCTION

COVID-19 has had an unprecedented impact on virtually every aspect of our lives, and youth sports are no exception. As groups across the country look to reinitiate youth sports, there are competing risks that should be considered within any specific setting. Physical activity and sport participation have tremendous physical and mental health benefits for children, but this must be balanced against the possibility of viral transmission during activity. Early research suggests that school and sport cancelations during the initial months of the COVID-19 pandemic were associated with significant decreases in physical activity and worsening of depressive symptoms in children and athletes.^5,6,9^ It has been projected that prolonged restriction could contribute significantly to long-term increases in obesity and mental health disorders.^3,8,12^ Together these results suggest that isolation and physical inactivity during COVID-19 restrictions may represent a significant threat to physical and mental health in children.

Nonetheless, COVID-19 continues to spread throughout the country and more than 200,000 deaths in the US have been attributed to the virus.^16^ In general, children and young athletes appear to experience milder symptoms than adults,^1^ but there is concern that sport participation could contribute to community transmission and expose individuals who may be more likely to experience severe consequences of the disease.^13^ Multiple groups have developed protocols and recommendations for sport participation to reduce this risk,^2,4,10,11,15^ but it remains unclear whether youth sport participation with risk mitigation procedures in place increases the incidence of COVID-19 among youth sport participants.

Soccer remains the most popular sport in the United States, and as local guidelines began to ease during the summer of 2020, many youth soccer organizations began to reinitiate participation. Recommendations have been published regarding procedures to mitigate COVID-19 risk within soccer specifically,^7,14^ but it is unknown whether this has resulted in an increased risk of COVID-19 among youth soccer participants, which could potentially contribute to a greater degree of community spread. Consequently, this limits discussions about whether the risks of COVID-19 transmission during youth sports participation would outweigh the physical and mental health risks of restriction from participation. Therefore, the purpose of this study was to identify the incidence of COVID-19 among a nationwide sample of youth soccer players, as well as the current mitigation procedures being implemented to reduce the risk of disease spread.

## METHODS

Through a collaboration with the Elite Clubs National League (ECNL), a leading nationwide youth soccer competition and development platform, surveys were distributed to the directors of all member clubs between August 26, 2020 and August 31, 2020. Soccer club directors were asked to provide information for their entire organization. In addition to facility location, clubs were asked whether they had reinitiated participation in soccer since the initial COVID-19 restrictions in the spring of 2020. Those clubs who reported reinitiating soccer were asked what stage of participation to which they had returned (“individual training only”, “non-contact, physically distanced”, “contact, not physically distanced”, or “other”), the date of their reinitiation, the total number of players and staff members participating, the number of trainings and games that their players had participated in, and the number of players and staff members who had been diagnosed with COVID-19 since restarting soccer participation. Clubs who reported a positive case of COVID-19 were asked to provide the number of cases among players or staff that were traced back to soccer participation, and the number that resulted in hospitalization or death. Finally, each club was asked whether they had a formal plan for COVID-19 risk reduction and which of 12 different procedures they utilized (see Table 4). Follow up was conducted with clubs regarding anyreporting outlying values.

**Table 1.** Comparison of COVID-19 cases, case rate, and incidence between youth soccer players from clubs that had reinitiated group soccer participation and US children nationwide.

|  | Youth Soccer Players | US Children<sup>a</sup> |
| --- | --- | --- |
| Total population | 85,861 | 75,471,700 |
| Total cases | 218 | 360,263 |
| Total person-days | 6,317,004 | 5,283,019,000 |
| Cases per 100,000 people | 254 | 477 |
| Incidence rate (cases / person-day)<sup>b</sup> | 0.0000345 | 0.0000682<sup>c</sup> |

<sup>a</sup>Data for 6/18/2020 to 8/27/2020, extracted from the American Academy of Pediatrics “Children and COVID-19: State Data Report”, August 27, 2020; <sup>b</sup>Incidence rate ratios calculated using unbiased median estimation and mid-p confidence intervals. <sup>c</sup>p<0.001 versus Youth Soccer.

**Table 2.** Comparison of COVID-19 cases, case rate, and incidence between youth soccer players and population data from each respondent county (where available).

|  | Youth Soccer Clubs<br>(n=94) | Respondent Counties<br>(n=56)<sup>a</sup> |
| --- | --- | --- |
| Total population | 58,588 | 72,654,438 |
| Total cases | 157 | 665,980 |
| Total person-days | 4,296,925 | 5,085,810,646 |
| Cases per 100,000 people | 268 | 917 |
| Incidence rate (cases / person-day)<sup>b</sup> | 0. 0000365 | 0.000131<sup>c</sup> |

<sup>a</sup>Data extracted from available public information from local health authority websites. <sup>b</sup>Incidence rate ratios calculated using unbiased median estimation and mid-p confidence intervals. <sup>c</sup>p<0.001 versus Youth Soccer.

**Table 3.**
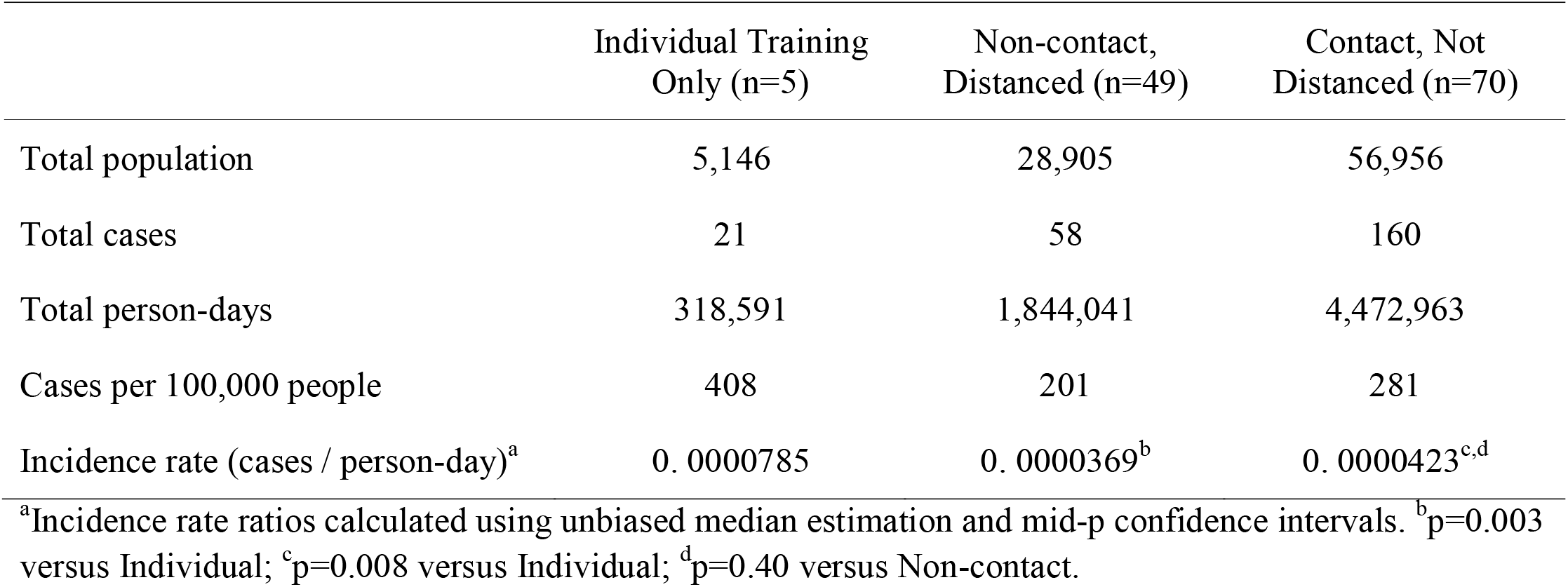
Comparison of COVID-19 cases, case rate, and incidence rate in youth soccer players between clubs at different phases of progression to return to soccer.

|  | Individual Training<br>Only (n=5) | Non-contact,<br>Distanced (n=49) | Contact, Not<br>Distanced (n=70) |
| --- | --- | --- | --- |
| Total population | 5,146 | 28,905 | 56,956 |
| Total cases | 21 | 58 | 160 |
| Total person-days | 318,591 | 1,844,041 | 4,472,963 |
| Cases per 100,000 people | 408 | 201 | 281 |
| Incidence rate (cases / person-day)<sup>a</sup> | 0. 0000785 | 0. 0000369<sup>b</sup> | 0. 0000423<sup>c,d</sup> |

<sup>a</sup>Incidence rate ratios calculated using unbiased median estimation and mid-p confidence intervals. <sup>b</sup>p=0.003 versus Individual; <sup>c</sup>p=0.008 versus Individual; <sup>d</sup>p=0.40 versus Non-contact.

**Table 4.** Association of local COVID-19 incidence and phase of soccer play with COVID-19 incidence among youth soccer clubs.^a^

| Variable | Estimate | Standard Deviation | p |
| --- | --- | --- | --- |
| Local (county) incidence | 5900 | 770 | <0.001 |
| Phase of play - non-contact, distanced | 0.73 | 0.84 | 0.38 |
| Phase of play - contact, not distanced | 0.57 | 0.82 | 0.49 |

<sup>a</sup>Estimate, standard deviation and significance level for each variable derived from a negative binomial regression model to predict COVID-19 cases within each club, including local COVID-19 incidence and phase of play (reference="Individual Training") as covariates as well as log(player-days) as an offset.

Data were initially evaluated using descriptive statistics including estimates of central tendency (mean, median) and variability (standard deviation, interquartile range, range) for continuous variables and counts and percentages for categorical variables. Duration since restarting was determined for each club as the number of days between the date of reinitiation and the date the survey was completed, and player-days for each club was calculated as the product of the duration and the number of players that had returned within the club. The overall case rate was expressed as the number of cases per 100,000 players (reported cases / total number of players * 100,000). The overall incidence rate was calculated as the total number of reported cases divided by the total player-days. Given a median duration of soccer participation of 73 days for the respondent clubs, the national pediatric case rate and incidence rate were determined over 10 weeks prior to the survey administration using data provided by the Academy of Pediatrics on August 27, 2020. Case rates between the total club data and national data were compared descriptively and incidence rates were compared using median unbiased estimation.

In addition, the case rate and incidence rate during the 10 weeks preceding survey responses were determined for each county in which a respondent club was located from publicly available online information from the local health authority. For those clubs with available county information, case rates and incidence rates were compared between the aggregate club and county data. Case and incidence rates were also compared between those clubs who had returned to “individual training only” (individual), “non-contact, physically distanced” (non-contact) play and “contact, not physically distanced” (contact) play. In both cases, case rates were presented for descriptive comparison, while incidence rates were compared using median unbiased estimation. Finally, to evaluate the relationship between COVID-19 cases and phase of play while adjusting for the local disease burden, a negative binomial regression model was developed to predict the number of club cases, including the incidence rate within each county in the prior 10 weeks, the phase of current soccer play for each club (contact, non-contact) and the log of player-days as an offset in the model. A large number of clubs reported 0 cases, but a separate zero-inflated negative binomial model with the same predictor and outcome variables yielded virtually identical results and model fit parameters. All statistical analyses were performed in R statistical software (The R Foundation for Statistical Computing).

## RESULTS

Surveys were completed by 129 respondents, of which 124 had restarted playing soccer since local restrictions were put in place at the beginning of the COVID-19 pandemic. These 124 respondents represented 91,007 players from 34 states who have participated in 45,574 trainings and 6,208 games since restarting. The time since restarting varied across clubs, with a median duration of 73 days (interquartile range (IQR): 53 - 83 days). With respect to phase of return, 5 clubs (4%) remained in individual training only, 49 clubs (39%) had progressed to non-contact / physically distanced group play, while 70 clubs (57%) had progressed to soccer participation that involved contact / unrestricted group play.

Two hundred eighty-two positive cases of COVID-19 were reported, including 239 players and 43 staff members from seventy-eight clubs (63%). One club initially reported an outlying value for player cases, which on follow up was determined to have been misinterpreted as the number of players who had been quarantined due to exposure, and subsequently clarified to include only players diagnosed with COVID-19. Among youth soccer players specifically, this represents 263 cases of COVID-19 per 100,000 children and an incidence rate of 4.3 × 10^−5^ cases per person-day.

Among the 119 clubs that had progressed to “non-contact” or “contact” group play, 218 cases were reported among players. The comparison of COVID-19 within youth soccer clubs and national pediatric data is shown in Table 1. Youth soccer players had a lower incidence rate than the general population of children in the United States (incidence rate ratio [IRR] = 0.511, 95% CI = [0.40-0.57], p<0.001). Population-level COVID-19 data was publicly available for 55 of the 76 counties in which respondent clubs were located. The comparison of COVID-19 within these clubs and their respective counties is shown in Table 2. Youth soccer participants had a lower incidence rate than the general population from the respondent counties (IRR = 0.202 [0.19-0.21], p<0.001).

The comparison of COVID-19 incidence by phase of return to soccer is shown in Table 3. Players engaging in non-contact group play had a lower incidence rate than those who remained in individual training (IRR = 0.47, 95% CI = [0.29-0.80], p=0.003), as did those in contact group play (IRR = 0.54, 95% CI = [0.35-0.88], p=0.008). No difference in incidence rate was identified between those groups is contact and non-contact group play (IRR = 1.10, 95% CI = [0.85-1.50], p=0.40). In the multivariable, negative binomial model adjusted for local COVID-19 incidence, the number of club cases was positively associated with the local incidence rate, but not associated with the phase of return (Table 4). Of the 282 cases reported, 1 case in a player was reportedly attributed to transmission during soccer. No cases were reported to have resulted in hospitalization or death. Of the clubs that had restarted playing soccer, 124 (100%) responded that they had a formal plan in place regarding COVID-19 risk reduction procedures. The median number of risk mitigation procedures utilized was 8 (interquartile range: 6-10), and the majority of clubs reported incorporating a number of different COVID-19 risk reduction procedures (Table 5).

**Table 5.** The proportion of youth soccer clubs that report implementation of different COVID-19 risk reduction procedures.

| Risk mitigation procedure | % of clubs |
| --- | --- |
| Player/staff symptom monitoring | 93% |
| Player/staff temperature checks at home | 85% |
| Player/staff temperature checks on site | 32% |
| Face mask use for players off the field | 80% |
| Face mask use for staff | 85% |
| Social distancing for players while playing | 54% |
| Social distancing for players off the field | 92% |
| Social distancing for staff | 85% |
| Increased facility disinfection | 74% |
| Staggered arrival and departure times for events | 84% |

## CONCLUSIONS

These findings suggest that the incidence of COVID-19 among youth soccer athletes after reinitiating participation during the summer of 2020 is relatively low. The number of cases per 100,000 players, and the incidence rates reported by the participant clubs since returning to play, are both lower than those reported by the American Academy of Pediatrics for children in the United States during the same time period. The disease burden and risk of COVID-19 varies considerably in different parts of the country, however, so we also sought to compare the COVID-19 incidence of the respondent clubs with the data from the county in which the club is located. We again found that this population of youth soccer athletes had lower case and incidence rates than the populations of the counties in which their clubs are located. While there are limitations in comparing the overall case and incidence rates in this study to the case and incidence rates from the general populations in each county, county-level data for children over the 10-week timeframe prior to the study was not publicly available for comparison in most cases. Nonetheless, our data does not appear to suggest that participation in youth soccer activities results in an increased risk of COVID-19 among participants. In addition, these findings agree with the existing literature regarding the rarity of severe outcomes of COVID-19 in children,^1^ as none of the cases were reported to result in hospitalization or death. Finally, of the 282 positive cases reported among players, only 1 was attributed to transmission during soccer activities.

As a secondary analysis, we sought to determine whether the phase of return to soccer was associated with the risk of COVID-19 among youth players. Soccer is an outdoor sport with relatively widely distributed players who do not appear to spend significant amounts of time within close proximity to each other during normal game play. Nonetheless, it has been generally suspected that contact participation would result in an increased risk of viral transmission. Initially we found that those clubs that remained in a phase of individual training only had higher incidence rates than clubs that had progressed to both non-contact and contact group play, with no difference between the non-contact and contact groups. Because local decisions regarding the progression of participation in youth sports is at least partly a function of the local COVID-19 disease burden, the higher incidence rate among clubs that had not progressed past individual training is likely simply a reflection of higher background incidence among those clubs. Indeed, the case and incidence rates among the clubs that remained in individual training were similar to those of children in the US in general.

After adjusting for the local incidence rate in the multivariable model, we did not find a significant relationship between phase of return and the number of COVID-19 cases within each club, while local COVID-19 incidence remained a significant predictor. It is difficult to fully interpret these results, as most of the clubs that have progressed to contact play likely progressed through a period of non-contact play initially, and we do not have data regarding whether the reported cases among these clubs occurred during the initial non-contact phase or later during the contact phase. Nonetheless, incidence rates were very similar between clubs participating in non-contact and contact group play, and neither non-contact nor contact group play was associated with an increased incidence of COVID-19 after adjusting for the local COVID-19 incidence. This may suggest that the COVID-19 risk among these youth soccer clubs is primarily a function of the background disease burden, rather than participation in soccer. Given the outdoor, physically distributed nature of soccer, it may be the case that participation in soccer activities does not result in a significant increase in COVID-19 risk when risk mitigation procedures are utilized.

Multiple sports organizations, academic organizations, research groups and federal and local health authorities have developed and recommended risk mitigation procedures to be incorporated by sports organizations during the reinitiation of participation.^2,7,10,11,14,15^ Within youth sports, these may vary between activities but generally include recommendations regarding social distancing, mask use, symptom monitoring, hygiene and disinfection practices, staggered arrival and departure times, and return to play procedures following diagnosis or exposure. Among the respondent clubs within this study, all reported having a formal COVID-19 plan in place, and the majority reported utilizing a broad range of risk mitigation procedures. For example, over 80% reported utilizing symptom monitoring and temperature checks among players and staff, social distancing and face mask use by staff and by players off the field, and staggered arrival and departure times. This suggests that youth soccer organizations are motivated to maintain an environment to reduce the risk of COVID-19 transmission during reinitiation. It should be recognized that this study cannot comment on the incidence or transmission risk of COVID-19 among attendees at soccer events other than players and staff. While this risk remains undefined, it nonetheless represents an important potential contribution to community COVID-19 spread, and a number of the risk mitigation procedures being implemented are also intended to help reduce the risk of infection among attendees. Consequently, these should remain a priority for youth sports organizations.

This survey-based study has several limitations. The information self-reported by the clubs cannot be directly verified through medical records or another independent source. It is unlikely that clubs were utilizing serial testing of asymptomatic players, so the true incidence among these athletes cannot be defined. Nonetheless, the testing of symptomatic or exposed youth soccer athletes is likely comparable to the indications for testing of children in the United States in general, making the comparison of the incidence among youth soccer players to the background case and incidence rates applicable. Similarly, we were only able to obtain data for the whole population of respondent counties during the 10 weeks prior to the study, and we are not able to compare our data to local, pediatric case or incidence rates. Formal contact tracing was likely not employed by the clubs involved, and we cannot directly account for the possibility of transmission between players that went unidentified. Similarly, we do not know how many cases were attributed to sources outside of soccer as opposed to having no source identified; therefore, we cannot address a true transmission rate within youth soccer activities. Finally, while this data represents information regarding a large number of male and female youth athletes from a nationwide sample, it should not be generalized to other sports.

In conclusion, these results suggest that the incidence of COVID-19 among youth soccer athletes is relatively low when compared to the background incidence among children across the country, and when compared to the incidence within the local general population. In addition, no difference in COVID-19 risk was identified between those clubs that had returned to contact participation and those that had not, even after adjusting for the local background disease burden and the number of risk reduction procedures in place. The youth soccer clubs in this study universally report having a formal plan in place to reduce the risk of COVID-19 spread and the overwhelming majority report using a variety of risk mitigation procedures. While we hope that this information will help contribute to the ongoing discussions about the relative risks and benefits of youth sport participation, we should recognize that COVID-19 risk will surely vary between sports and across different areas of the country. In addition, this study cannot address the potential risk of transmission among attendees at youth sporting events, which remains a vital area of future research. Therefore, this data should represent an initial step toward developing a more complete picture of the relative risk of COVID-19 transmission during sport participation for children and there remains an urgent need to expand these efforts in order to make informed decisions within specific contexts.

## Data Availability

Requests for access to the data referred to in the manuscript can be directed to the primary author.

## ACKNOWLEDGEMENTS

There are no funding sources to report for this study. Dr. Watson serves as the Chief Medical Advisor for the Elite Clubs National League and is supported by grants from the National Center for Advancing Translational Sciences (UL1TR002373; KL2TR002374). We are grateful for the resources and support of the UW Institute for Clinical and Translational Research. There are no other relevant conflicts of interest to disclose.

## Notes

### Funding Statement

None of the authors or their institutions received any funding for any aspect of the submitted work.

### Author Declarations

The Health Sciences IRB for the University of Wisconsin determined that this research did not require formal IRB review because it did not involve human subjects as defined by federal regulations.

